# Can we Generate and Validate a Pregnancy Register for Research in the UK Greater Manchester Care Record Database Using Routine Primary Care Records

**DOI:** 10.64898/2026.09.30.26364393

**Authors:** Chantelle Cornett, George Tilston, Glen P. Martin, Victoria Palin

## Abstract

**Background:** The Greater Manchester Care Record (GMCR) integrates health and social care data for 2.8 million individuals. While it contains maternity information, identifying individual pregnancies, including timing and outcomes, is complex due to variations in healthcare interactions and recording practices. This study develops open-source code to create a pregnancy register within the GMCR using diagnosis- and procedure-based algorithm established in other databases.

**Methods:** 1,448 maternity SNOMED clinical codes identified pregnancy-related primary care records among women aged 14–49 years between March 2012 and August 2023. Codes were grouped by pregnancy outcome, with a hierarchical ruling approach where multiple outcomes appeared within one episode. Pregnancy start dates were derived from hierarchical coded events or imputed using biologically plausible duration based on outcome type. Internal validation compared GMCR register episodes against Hospital Episode Statistics (HES) hospital maternity admissions; external validation compared outcome rates against published UK estimates.

**Results:** The algorithm was developed across 379,832 women, identifying 880,926 pregnancies. Internal validation showed nearly half of register deliveries had a corresponding HES record, with a positive predictive value (PPV) of 48.1%and a median date difference of 17 days (IQR 1–30). External comparison revealed live birth (927.8 vs. 995.6 per 1,000 deliveries) and stillbirth (2.12 vs. 3.9 per 1,000 births) rates close to national estimates but a notable discrepancy for miscarriage (8.00 vs. 15.3 per 100 pregnancies) and termination of pregnancy (5.36 vs. 25.1 per 1,000 women). Demographic alignment was strong for maternal age (31 vs. 29.8 years) and deprivation (IMD10 median: 3 vs. 2.6), though ethnic minority representation was expectedly higher in the GMCR (37.0% vs. 28.7%).

**Conclusions:** This pregnancy register enriches the GMCR for maternal health research, enabling more comprehensive and regionally tailored studies to inform healthcare delivery and policy. This work represents an important first step in developing a scalable, open-source algorithmic tool to support pregnancy research across diverse populations. By making the code openly available, it can be adapted for use in federated data environments, enhancing reproducibility and collaboration.

## Introduction

Identifying and tracking individual pregnancies within routine healthcare records is inherently challenging. Maternity care in the UK is often delivered across multiple NHS sites, and variations in how care interactions are recorded make it difficult to consistently identify pregnancies within research patient records, hindering efforts to assess maternal and fetal health risk, monitor outcomes, and ensure equitable care. Various methods have been implemented to identify pregnancy episodes, such as rule-based backward-looking algorithms anchored to outcomes codes or forward looking logic from early pregnancy markers, [1]. Approaches in the UK focus on primary-care and Hospital Episode Statistics (HES) data [2, 3], including recently published work using the QResearch database [4].

Across the UK, there is a growing policy drive towards integrated care systems and shared care records that link primary care, secondary care, and social care data to support research and service planning at scale. The Greater Manchester Care Record (GMCR) is an early example of this type of regional, integrated dataset, first established in response to the COVID-19 pandemic to bring together essential information on its 2.8 million citizens [5]. Greater Manchester has a culturally and socioeconomically diverse population, with the latest census (2021) estimating that 28.7% of Greater Manchester’s residents are from an ethnic minority (above the national average of 26.5%) [6]. In addition, 34.2% of the Greater Manchester population are within the lowest National Statistics Socio-economic Classification (NS SEC) group (NS SEC 6-8), compared to 30.6% nationally [7]. This diversity, combined with the breadth of linked health and social care data available through the GMCR, makes Greater Manchester a useful setting in which to develop and test pregnancy identification methods that could subsequently be applied across other integrated care records nationally.

The aim of this paper was to develop an open-source algorithm to generate a pregnancy register using the GM Care Record, building on similar methods successfully implemented in other parts of the UK [2, 3]. By making the code openly available, the approach is designed to be adapted across other integrated care datasets, supporting federated analysis, reproducibility, and the growing shift towards regionally linked data infrastructures for pregnancy research.

In this paper, we outline an algorithm adapted from the CPRD GOLD pregnancy registry, [2] reflecting the methodological approaches available at the time of development (2022-2023) and implemented within an open-source, adaptable framework. The methods follow the approach and validation described by Minassian et al, with minor adaptations detailed. We also describe the resulting output population from the GMCR pregnancy registry and discuss the strengths and limitations of this approach, enabling users to assess its suitability for pregnancy health research and its potential applicability to other regional data environments.

## Methods

### Data Sources and Study Population

We used GMCR primary care data, secondary care Hospital Episode Statistics (HES) admissions records for maternity-related admissions, and open-source data to compare the GMCR pregnancy register statistics to for validation. We included all primary care records for female patients, aged 14-49, with a pregnancy-related SNOMED code between the dates of March 1st, 2012, to August 31st, 2023, who are recorded in the GMCR. For included patients, we extracted key demographic information when recorded. No further restrictions were applied.

### Code Lists

At the time of data curation clinical code sets from the established CPRD GOLD algorithm [2] were used to identify pregnancy episodes within the GMCR cohort. As the code set used Read codes, a clinical terminology in place prior to the national transition to SNOMED CT, the Read codes were converted to SNOMED CT clinical codes by the data provider for the GMCR. A full set of clinical codes used are available via the GitHub Repository [8]. Following the CPRD GOLD approach, codes were then grouped into pregnancy related concepts, where codes could belong to multiple concepts (Table 1).

**Table 1.**
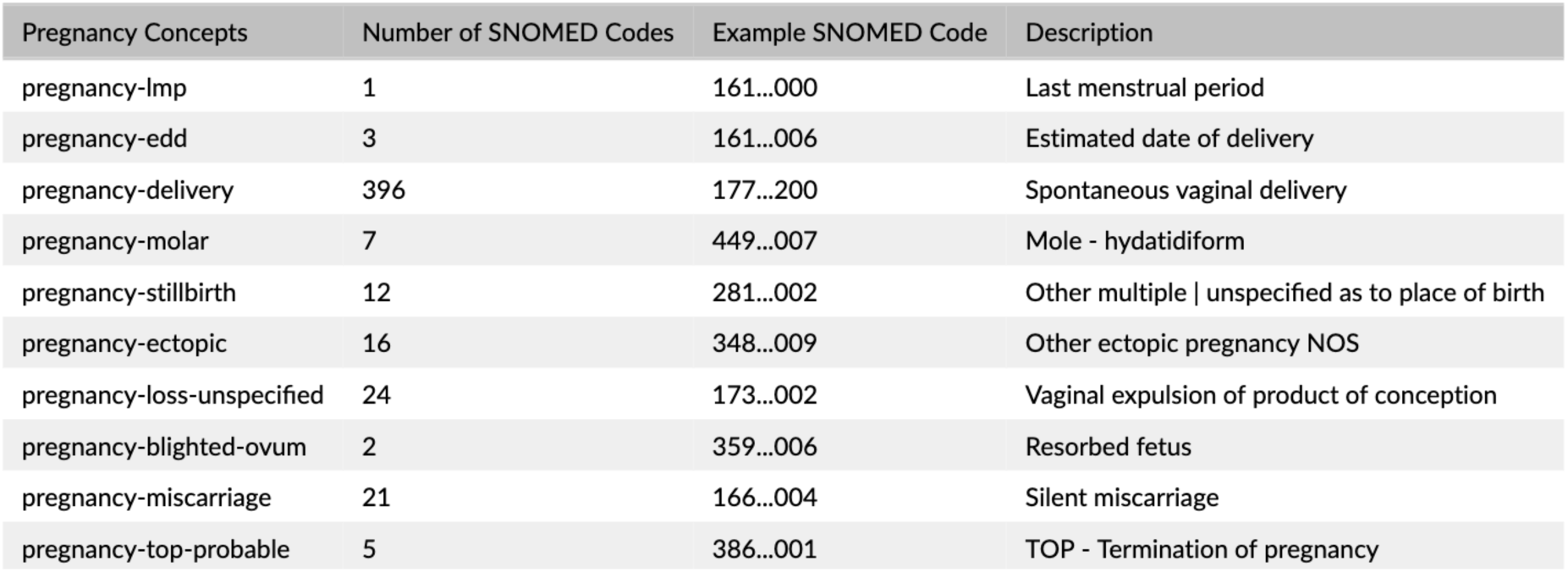
Pregnancy concepts and SNOMED codes with examples. (Abbreviations: NOS - Not otherwise specified).

### Algorithmic Approach to Create the GMCR Registry

The algorithm below was adapted from the approach described by Minassian et al. [4]; points at which the GMCR implementation departs from the original approach are flagged in the text below and summarised in Figure 1.

**Figure 1.**
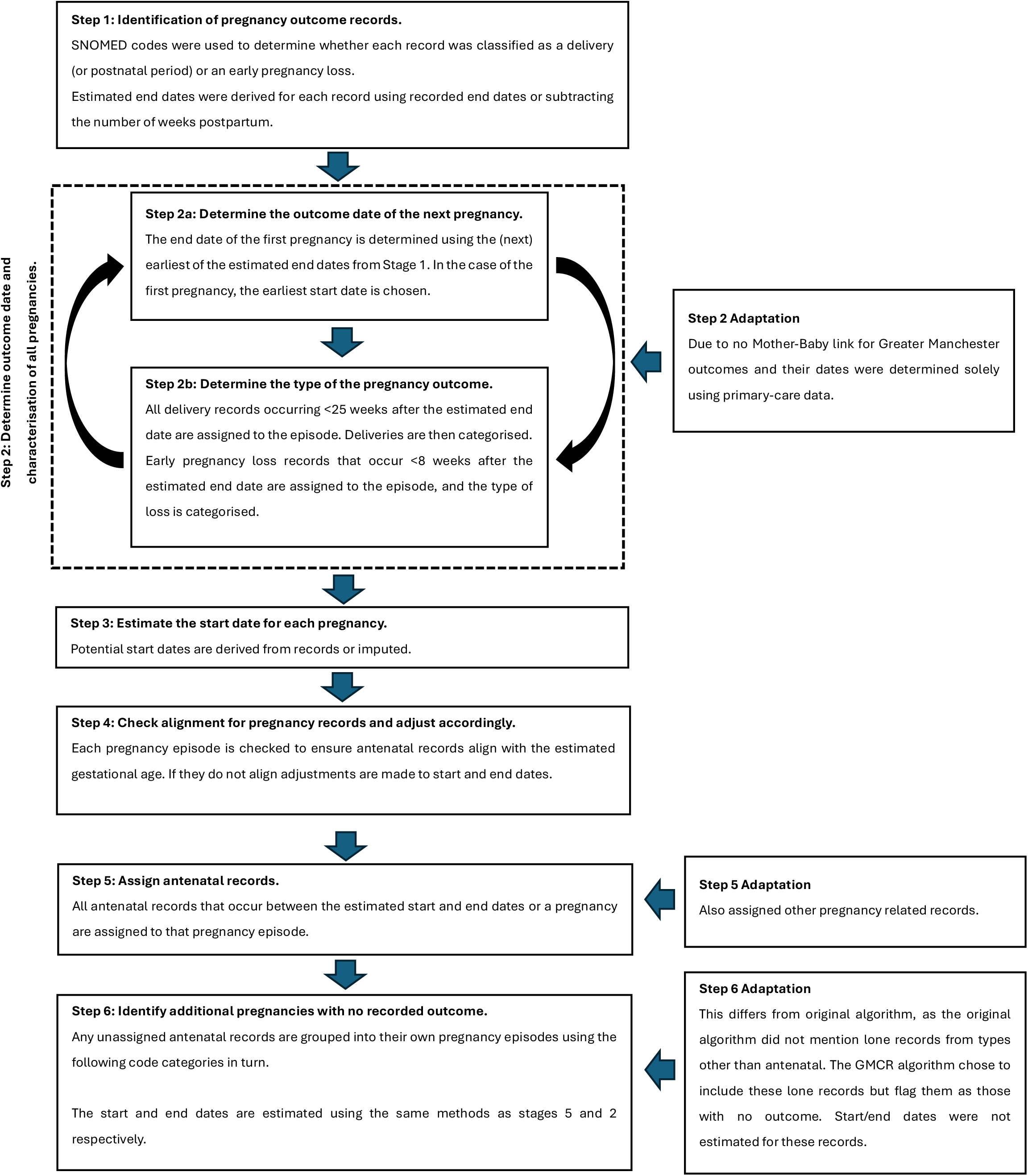
A flow diagram showing the adapted stages in the algorithm based on the Minassian et al.[2] approach.

The algorithm started by estimating pregnancy end dates for each patient using coded clinical events that explicitly indicated a pregnancy outcome. End-dates were therefore derived from clinically coded concepts, with delivery and pregnancy loss events identified and processed separately from the outset (all steps of the algorithm and their adaptations can also be viewed in Figure 1). Distinct pregnancy episodes were then constructed by grouping records relating to the same outcome type (delivery or loss) for each patient into distinct timeframes. For delivery outcomes, the first pregnancy episode was defined using the earliest recorded delivery-related date, and all subsequent codes recorded less than 25 weeks after this were assigned to the first episode. A minimum separation of 25 weeks was then applied before searching for each successive delivery episode on a record. For codes relating to early pregnancy loss, all records that occur less than 8 weeks after the first estimated loss date were assigned to that episode, allowing for a minimum separation of 8 weeks between successive losses. As there is no Mother-Baby link within the GMCR, outcome dates and characterisation were determined solely using primary care data, in contrast to the original CPRD GOLD approach.

After records were grouped into distinct pregnancy episodes, each episode’s outcome was assigned by matching the associated SNOMED CT code to the main outcome concepts (Table 1). For deliveries, outcomes were labelled as preterm, post-term, multiple or stillbirth where available. For pregnancy losses, the type of loss was determined by the assigned codes from the categories in Table 2. If more than one category occurred within the defined episode for each unique patient ID, the outcome was assigned based on the hierarchy in Table 2 (as used by Minassian et al. [2]).

**Table 2.** Hierarchy of pregnancy loss records, used for determining outcome type.

| Hierarchy | Early Pregnancy Loss Category |
| --- | --- |
| 1 | Ectopic |
| 2 | Termination of pregnancy |
| 3 | Miscarriage |
| 4 | Probable termination of pregnancy |
| 5 | Molar pregnancy |
| 6 | Unspecified pregnancy loss |
| 7 | Blighted ovum |

Once the records were grouped by pregnancy end date, the start date for each episode was estimated using date specific SNOMED CT codes if present within the episode cluster. Relevant records included codes for estimated date of delivery (EDD), estimated date of conception (EDC) or last menstrual period (LMP). For deliveries, the following order of priority was used when more than one type was present within a cluster to estimate the start date: (a) EDD subtracting 280 days from the entry date, (b) EDC subtracting 14 days, and (c) LMP where the event date was used. If the records indicated a delivery was pre-term, post-term, or multiple and if the records for EDD, EDC, and LMP were not available then these were imputed using biologically plausible estimates (see Table 3). Codes relating to postnatal or pregnancy loss counselling or postnatal without a specified time were not used to determine start dates due to the uncertainty surrounding how long they occur after a delivery or loss. Codes relating to incomplete or failed termination of pregnancy were also not used to determine start dates due to the uncertainty surrounding the timing of the event and pregnancy outcome.

**Table 3.** Default durations to impute the start date where necessary.

| Pregnancy Outcome | Default Duration (completed weeks) |
| --- | --- |
| Preterm delivery | 36 |
| Post-term delivery | 41 |
| Multiple delivery | 37 |
| All other deliveries | 40 |
| Ectopic pregnancy | 9 |
| All other pregnancy losses | 12 |

All episodes were then examined to ensure the length of each pregnancy was plausible. Gestational length was calculated for each pregnancy using the start and end dates. For post-term and term pregnancies, if the period of gestation was greater than 42 weeks then the end date was adjusted to 42 weeks from the start date. For preterm deliveries, if the gestation was greater than 36 weeks plus 6 days then the end date was adjusted to be 36 +6 from the start date. For multiple deliveries, if the gestation was greater than 37 weeks then the start date, then the end date is adjusted to 37 weeks from the start date. All pregnancies that had their start or end dates adjusted were flagged. Delivery and loss episodes were constructed independently for each woman, and following Minassian et al. [4] where the estimated dates of a delivery episode and a loss episode for the same woman overlapped, (e.g., a loss episode nested within a later delivery episode, potentially reflecting a threatened miscarriage that culminated in a live birth) the overlapping episodes were flagged as a “conflict” rather than resolved, leaving these episodes available for users to handle according to their own study requirements. All antenatal records occurring between the estimated start and end dates of a pregnancy were assigned to that episode. Any unassigned antenatal records that had no clear corresponding outcome (see Table 5 for all included outcomes and their frequencies) were flagged. Additionally, unlike the original Minassian et al. algorithm, which did not consider lone records from code categories other than antenatal codes, the GMCR implementation grouped lone, non-antenatal records into their own pregnancy episodes and flagged them as having no recorded outcome, rather than excluding them; start and end dates were not estimated for these episodes.

The final stage was to merge additional individual-specific variables to optimise the capabilities of the register. This included the Clinical Commissioning Group (CCG) of the primary care practice the patient was registered with, if a patient left a practice (de-registration or death) the exit date, and additional demographics including, year of birth, sex, ethnicity, Lower Layer Super Output Area (LSOA) code, Index of Multiple Deprivation (IMD) decide, Body mass index (BMI), date of BMI recording, current and worst smoking status, death within 28 days of a positive COVID-19 test, death within 28 days of a positive COVID-19 confirmed, and death date.

The final pregnancy register contains and describes all pregnancies recorded in the GMCR with one row per pregnancy. Woman may have more than one pregnancy episode. An R script has been made available as an open-source tool in GitHub for further development or adaptation [9].

## Validation Methods

### Internal Validation

To assess the validity of the algorithm we compared the dates for delivery outcomes in the pregnancy register to the secondary Admitted Patient Care (APC) HES database, which contains records of all inpatient admissions to NHS hospitals in England. The data contained admissions spells between January 1^st^, 2018, and March 1^st^, 2022, for women aged 14 to 49 years registered with a GMCR primary care practice during this period. Agreement between admission spells relating to a birth event recorded in HES APC and those documented in the GMCR Register were examined by comparing start and end dates. A delivery was considered matched across the two sources if the pregnancy outcome dates were within 12 weeks of each other. This time window accounts for possible inaccuracies in the Register’s date estimations and accommodates known delays in updating records when information moves from secondary to primary care settings. Using a broader matching period helps reduce the risk of misclassifying the same pregnancy as separate events. Using HES admissions as the reference standard, we evaluated the estimated PPV against HES recorded births as well as the completeness and temporal accuracy of pregnancy outcome recordings in the Register. For deliveries appearing in both sources and meeting the 12-week criterion, we also examined agreement on gestational age in completed weeks.

To understand factors contributing to unmatched cases, we carried out several sensitivity analyses. First, recognising the lag in GP-recorded hospital deliveries, and having a wider date coverage for the GM registry than the HES data, we included all Register-recorded pregnancies in the 6-month period after the HES follow up ended and excluded all Register-recorded deliveries in the first 6 months of the HES follow up period. This approach, like that applied by Minassian et al. [2], aims to avoid underestimating agreement due to delayed data entry. We also explored how the presence of repeated pregnancy and postpartum condition codes influenced the number of pregnancies identified and their concordance with maternity admission records. Finally, we compared demographic characteristics of the population in the GMCR pregnancy register with those of the broader Greater Manchester population to evaluate representativeness and alignment using data from the Office for National Statistics (ONS) and other published sources. To enable these comparisons, we defined three separate cohorts corresponding to the years 2015, 2021, and 2022.

### External Validation

The aim of the external validation was to assess the accuracy of the Register’s estimated rates of outcomes by comparing them with published estimates. [10–13]

As published estimates for these outcomes related to different reference years, each outcome rate was estimated for the specific year matching the corresponding published comparator, as detailed below. We calculated live birth and termination rates for 2022, matching the reference year of the relevant ONS and DHSC publications. [10, 11] The live birth rate was defined as the number of live birth deliveries and stillbirths per 1,000 births, while the termination rate was defined as the number of terminations per 1,000 women aligning with the units of measure used by the published estimates. To improve capture, we broadened the definition of termination to include probable termination and unspecified loss (see Table 5 for example codes). Miscarriage rates were estimated for 2021 and defined as the number of miscarriages divided by the total number of pregnancies with known outcomes. As with terminations, we expanded the miscarriage definition to include cases coded as unspecified loss. For 2015, we estimated the prematurity rate, matching the year of the relevant ONS Birth Characteristics dataset [13], defined as the number of preterm deliveries (less than 37 weeks of gestation) per 1,000 births (including both live births and stillbirths). This included estimating preterm births by using the prematurity flag in the Register, based on relevant SNOMED codes. We also broadened this definition to include additional deliveries with an estimated gestational age of less than 259 days (under 37 weeks).

## Results

### Register Composition and Recorded Outcomes

Across the study period, the GMCR Pregnancy Register identified 880,926 pregnancies among 379,832 women. These episodes comprised 518,539 deliveries and 58,071 early pregnancy losses, alongside 309,911 pregnancies with no recorded outcome. Of the pregnancies with unknown outcome, 10.54% were flagged as potentially ongoing at the end of follow-up (based on the earliest antenatal record occurring <38 weeks before the patient left the practice or the end of the study period). The distribution of pregnancies per woman and by outcome is presented in Table 4 and Table 5, respectively. Pregnancy history per woman indicated that repeated pregnancies were common: 36.30% of women had one pregnancy recorded, 22.15% had two, and 14.52% had three; the median was two pregnancies (IQR, 1–4).

**Table 4:** Number of pregnancies per woman.

| **Number of Pregnancies (n)** | **Women with n Pregnancies, N (%)** |
| --- | --- |
| 1 | 137 868 (36.30) |
| 2 | 84 121 (22.15) |
| 3 | 55 159 (14.52) |
| 4 | 36 053 (9.49) |
| 5 | 23 559 (6.20) |
| 6 | 15 207 (4.00) |
| 7 | 9 511 (2.50) |
| 8 | 6 136 (1.62) |
| 9 | 4 056 (1.07) |
| 10+ | 8 162 (2.15) |
| Total women | 379 832 (100) |

**Table 5:** Pregnancy outcome, frequency and percentage.

| **Pregnancy Outcome** | **Frequency (%)** |
| --- | --- |
| Live birth | 469 219 (53.26) |
| Stillbirth | 1 066 (0.12) |
| Live birth and stillbirth | 11 429 (1.30) |
| Miscarriage | 49 195 (5.58) |
| Termination (TOP) | 418 (0.05) |
| Ectopic pregnancy | 5 832 (0.66) |
| Molar pregnancy | 985 (0.11) |
| Blighted ovum | 46 (0.01) |
| Unspecified loss | 1 590 (0.18) |
| Delivery based on a third trimester code | 36 399 (4.13) |
| Delivery based on a late pregnancy code | 426 (0.05) |
| Outcome unknown | 309 911 (35.18) |
| Total pregnancies | 880 926 (100) |

Outcome coding was predominantly composed of delivery-related episodes, with 469,219 pregnancies classified as live birth (53.3% of all pregnancies) and 1,066 as stillbirth (0.1%); an additional 11,429 were coded as both live birth and stillbirth (1.3%). Early loss outcomes included 49,195 miscarriages (5.6%), 5,832 ectopic pregnancies (0.7%), and smaller numbers of molar pregnancy, blighted ovum, and unspecified loss. A further subset of episodes were classified as delivery based on late-pregnancy evidence (e.g., third-trimester or late pregnancy codes, 4.2%) when a definitive delivery outcome code was absent. Among deliveries, the algorithm classified 25.33% as preterm and 24.98% as post-term, with 4.24% identified as multiple births. 35.2% of pregnancies had no known outcome.

Comparing demographics (Table *6*, Table 7) in the GM register to published estimates for Greater Manchester, most are in alignment [14], with the median IMD for women in the register being 3 (IQR: 1,6) compared to external estimates of the 10 authorities within Greater Manchester of 2.6. The average maternal age in the register was 31 years (IQR: 26, 35), agreeing with the external estimate for average maternal age in Greater Manchester was found to be 29.8 years. Compared to the register (37.04%) external sources estimate lower frequencies of ethnic minorities (28.7% of the total population [15]). Greater Manchester specific estimates could not be found for the other demographics.

**Table 6:**
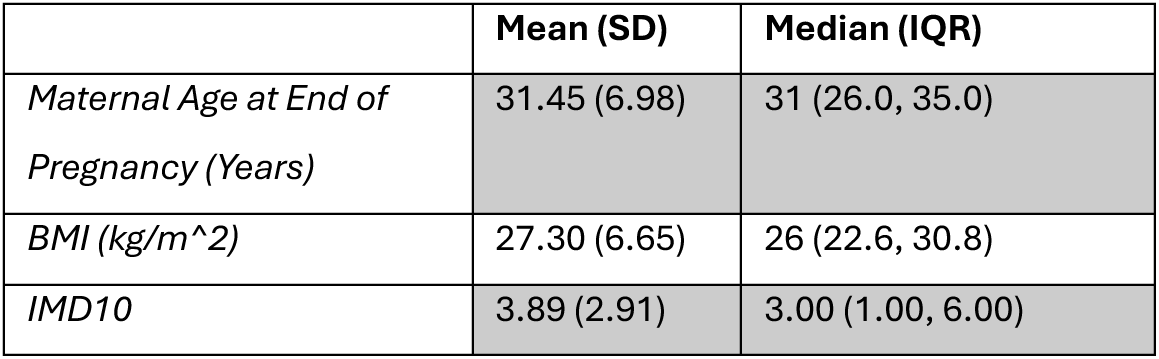
Demographic summary of maternal age at end of pregnancy (years), latest BMI (kg/m^2), and Index of Multiple Deprivation (IMD10).

**Table 7:**
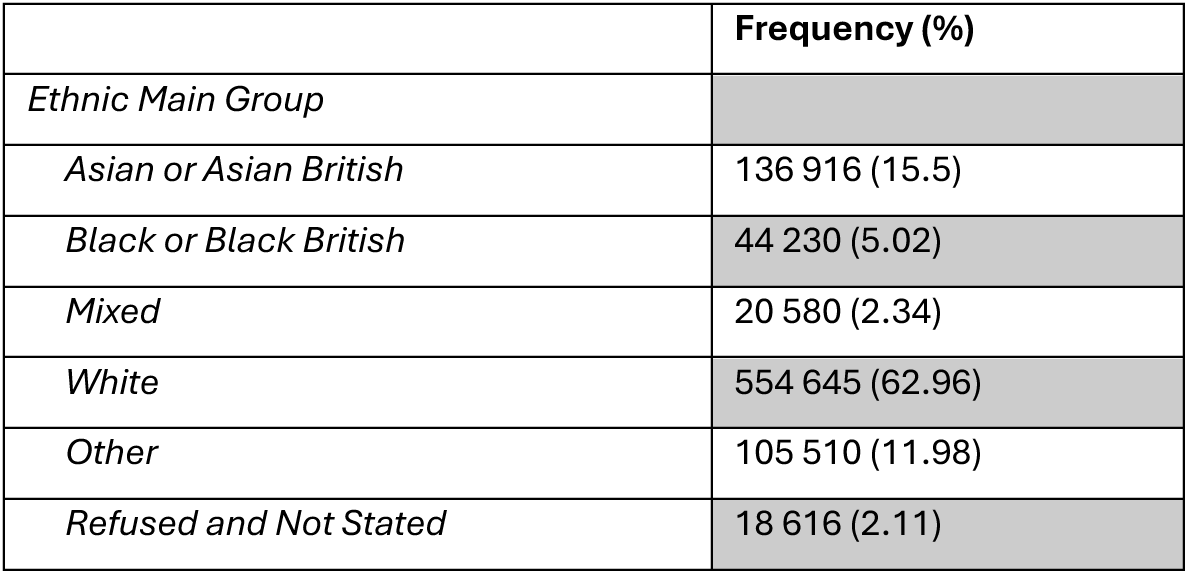
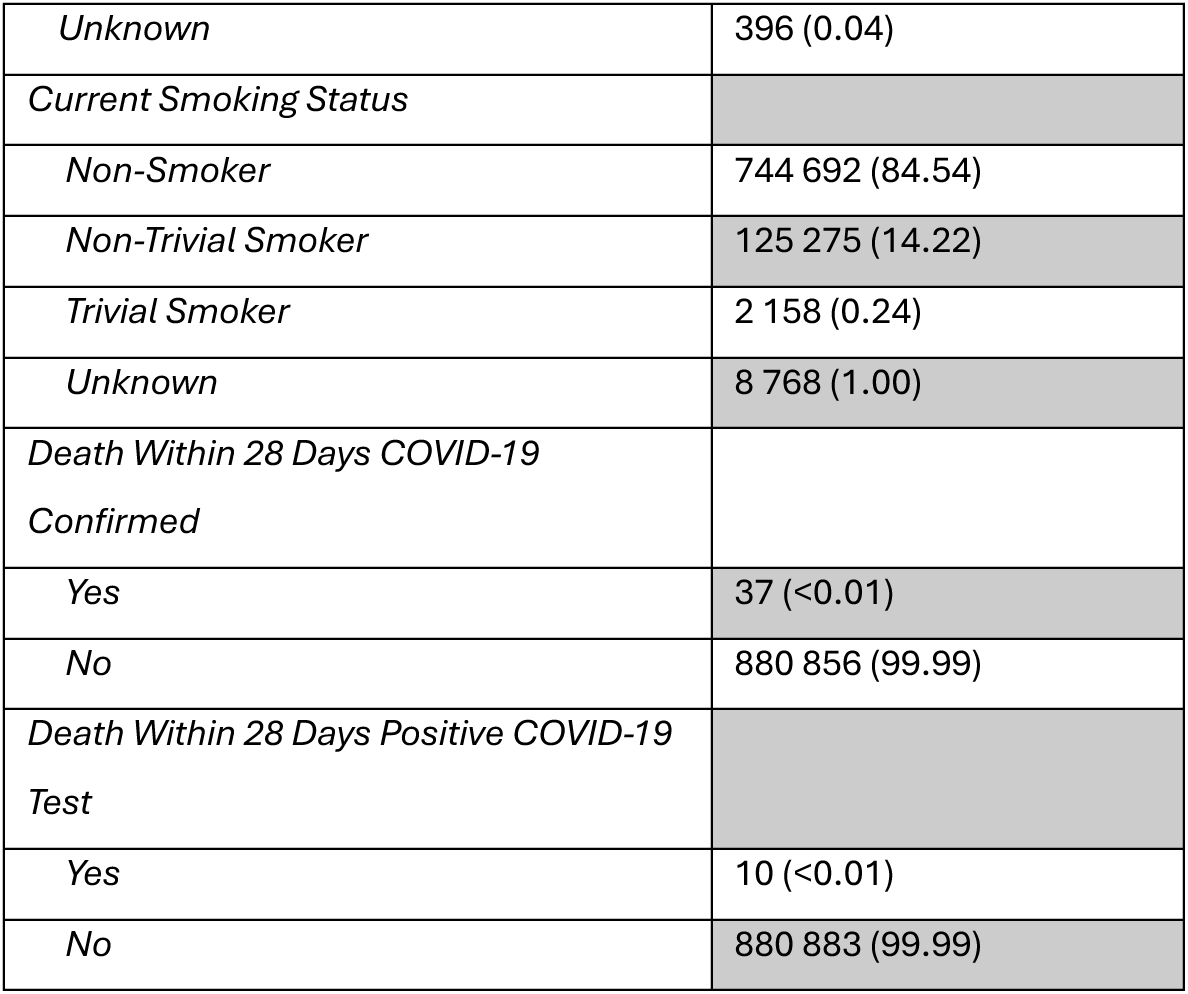
Demographic summary of ethnic main group, current smoking status, death within 28 days COVID-19 confirmed, and death within 28 days positive COVID-19 test.

|  | Frequency (%) |
| --- | --- |
| Ethnic Main Group |  |
| Asian or Asian British | 136 916 (15.5) |
| Black or Black British | 44 230 (5.02) |
| Mixed | 20 580 (2.34) |
| White | 554 645 (62.96) |
| Other | 105 510 (11.98) |
| Refused and Not Stated | 18 616 (2.11) |
| *Unknown* | 396 (0.04) |
| *Current Smoking Status* |  |
| *Non-Smoker* | 744 692 (84.54) |
| *Non-Trivial Smoker* | 125 275 (14.22) |
| *Trivial Smoker* | 2 158 (0.24) |
| *Unknown* | 8 768 (1.00) |
| *Death Within 28 Days COVID-19 Confirmed* |  |
| *Yes* | 37 (<0.01) |
| *No* | 880 856 (99.99) |
| *Death Within 28 Days Positive COVID-19 Test* |  |
| *Yes* | 10 (<0.01) |
| *No* | 880 883 (99.99) |

Comparing live-birth rates, the rate for the pre-pandemic period was significantly lower at 897.2 per 1000 births (CI: 894.0, 900.0) between January 2012 and December 2019, compared to 920.3 per 1000 births (CI: 914.0, 927.0) between January 2020 and December 2021 (COVID-19 pandemic). Live birth rates continued to be significantly higher post-pandemic at 925.3 per 1000 births (CI: 918.0, 932.0) from January 2022 to August 2023.

## Validation

### Internal Validation against maternity admissions

In the internal validation cohort (restricting to 2018-2022), 145,309 women had at least one delivery recorded in either data source: 190,635 deliveries were identified in the Register and 147,872 in maternity admissions (HES APC). Using a 12-week matching window, 91,645 deliveries were matched across sources in 52,799 women, representing 36% of the delivery cohort (Table 8). Using maternity admissions as the reference, approximately half of Register-recorded deliveries had a corresponding HES record (potential PPV 48.1%). Conversely, most HES deliveries were also present in the Register (sensitivity 62.0%). Unmatched episodes occurred in both directions: 51.9% of Register deliveries (98,990) had no HES match and 38.0% of HES deliveries (56,277) had no Register match. Among matched deliveries, 28.5% fell within the HES admission period and 54.5% differed by no more than three weeks. The median difference in delivery date (Register minus HES) was 17 days (IQR 1 to 30).

**Table 8.**
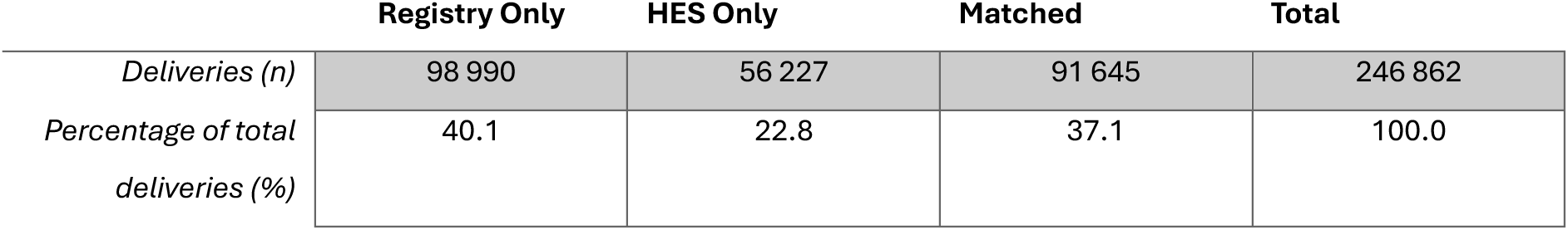
Internal validation: overlap between pregnancy registry deliveries and HES APC maternity admission (2018-2022)

Adjusting the observation window to account for data-availability and recording delays led to modest improvements. Excluding 51,485 Register deliveries occurring within the first 6 months of the HES follow-up period increased the potential PPV to 50.8%. Including 12,377 Register deliveries recorded within 12 weeks after HES follow-up ended increased sensitivity to 69.6%. Removing deliveries with no recorded start date, end date, or any antenatal/postnatal evidence excluded 4,977 deliveries.

### External Validation against published estimates

Table 9 presents the results of our external validation, comparing pregnancy outcome rates from the Pregnancy Register with estimates from the Office for National Statistics (ONS) and other published sources. For 2022, the Register identified 40,799 live births, 96 singleton stillbirths, and less than 20 terminations (or 170 when broadening the definition to include “probable terminations” and “unspecified losses”) among 31,620 women. Corresponding rates were lower than external estimates: 927.8 live births per 1,000 births (95% CI 919.0–937.0) versus 995.6 [10]; and 2.12 stillbirths per 1,000 births (CI 1.69–2.55) versus 4.4 [10]; and 0.38 terminations per 1,000 women (CI 0.17–0.60) versus 25.1. [11]

**Table 9.** GMCR Registry and external sources comparison. ONS live and stillbirth and DHSC statistics are specific to the Greater Manchester region. ONS – Office for National Statistics; DHSC – Department of Health and Social Care.

| **Pregnancy Outcome, Calendar Period** | **GMCR Registry** | **External Comparison Rate (Data Source)** |
| --- | --- | --- |
| Live birth rate, 2022 (per 1000 births) | 927.8 (919.0, 937.0) | 995.6 (Office for National Statistics; ONS)[10] |
| Stillbirth rate, 2022 (per 1000 births) | 2.12 (1.69, 2.55) | 4.4 (ONS)[10] |
| Miscarriage rate 2021 (per 100 pregnancies) | 7.76 (7.52, 8.00) | 15.3 (Sands) [12] |
| Miscarriage and unspecified loss 2021 | 8.00 (7.75, 8.24) |  |
| Termination rate, 2022<br>TOP only (per 1000 women) | 0.38 (0.17, 0.60) | 25.1 (DHSC) [11] |
| TOP and possible TOP | 0.38 (0.17, 0.60) |  |
| TOP, possible TOP and unspecified loss | 5.36 (4.55, 6.17) |  |
| Prematurity rate, 2015 (preterm only) (per 100 live births) | 3.72 (3.50, 3.90) | 7.64 (ONS) [13] |
| Prematurity rate, 2015 (preterm flag and gestational age < 259 days) | 13.20 (12.90, 13.50) |  |

For 2021, there were 50,596 pregnancies with known outcomes, including 4,007 miscarriages (4,137 when including unspecified losses). This corresponded to a miscarriage rate of 7.76%–8.00%, lower than the external estimate of 15.3% [12]. For 2015, the Register contained 45,674 deliveries. 1,702 were identified as preterm using the prematurity flag, and an additional 4,333 were identified using estimated gestational age, yielding an overall prematurity estimate of approximately 13.2%, compared with an ONS estimate of 7.64%.

## Discussion

### Summary of Main Findings

We developed the Greater Manchester Pregnancy Registry by adapting a previously published pregnancy identification algorithm and applying it within a large, federated primary care dataset. The final registry comprised 880,926 pregnancies among 379,832 women. Internal validation demonstrated moderate agreement with hospital delivery records. External comparisons suggested lower-than-expected rates for outcomes such as miscarriage and termination, alongside rates that were more closely aligned with external estimates for live births and stillbirths. Overall, the identified pregnancy cohort using our algorithm was demographically diverse and appeared broadly representative of the Greater Manchester region. This suggests this is the first step in creating a robust way to identify pregnancy episodes in routine primary care records for regionally tailored studies on maternal health to inform healthcare delivery and health policy. Beyond its immediate regional utility, the registry demonstrates that reproducible, modular algorithms can be deployed across federated primary care datasets this is of increasing relevance as national data infrastructure evolves toward integrated care system-level analysis.

### Comparison with the Literature

The lower observed rates of miscarriage and termination are consistent with under-recording in routine primary care data, particularly for outcomes that may be managed in secondary care, specialised services, or outside healthcare settings [16]. This pattern is consistent with limitations reported in CPRD-based pregnancy registries [2, 3]. In contrast, the comparatively high delivery estimate relative to national benchmarks was less anticipated. One plausible explanation is overestimation arising from imprecise gestational dating codes and episode date adjustments, where uncertainty in start dates and gestational age can lead to misclassification of term status. Additionally, the pattern of a rising live birth rate within the register contrasts with the national trend, where the total fertility rate for England and Wales has continued to decline since 2010 and reached a record low of 1.41 children per woman in 2024 [17]. Collectively, these findings reinforce existing challenges in using coded clinical data alone for pregnancy outcome identification, [2, 3, 16] while also illustrating that structured episode-based algorithms can recover meaningful population-level patterns when cross-checked against internal and external comparators.

### Data Quality and Availability

Our findings emphasise the dependence of pregnancy episode construction on the completeness and timing of coded events in primary care. In particular, early outcomes are vulnerable to incomplete capture, partly reflecting the well-documented challenge that hospital-managed events flow back to primary care primarily via discharge summaries rather than structured coded records. [2, 3, 16] Gestational timing can also be uncertain when key fields such as estimated delivery date are missing, recorded late, or inconsistently represented across care settings. These limitations have direct implications for study design, where analyses requiring accurate trimester assignment, gestation-dependent exposures, or precise interpregnancy intervals may need additional restrictions to higher-confidence episodes and explicit handling of date uncertainty. Looking ahead, integration of the Maternity Services Data Set and the NHS 10 Year Health Plan’s commitment to a single patient record rolled out first in maternity care, with universal digital care records by 2028, offer the potential to materially reduce these limitations over time.

### Cohort Demographics and Representativeness

A strength of the registry is the size and diversity of the cohort, which supports investigations across demographic strata that are often underpowered in smaller datasets. The demographic composition appears broadly consistent with regional estimates for Greater Manchester, [6, 7, 15] supporting the utility of the registry for locally relevant maternal and perinatal research, including equity-focused analyses.

### Internal and External Validation: Interpretation and Implications

Internal validation against hospital delivery records indicated only moderate concordance, with incomplete overlap between sources. This likely reflects a combination of differences in capture mechanisms and the downstream flow of information from maternity services into primary care records, rather than a single comprehensive source. External validation further contextualised these results where delivery-related outcomes were comparatively robust, whereas early pregnancy losses and terminations showed larger discrepancies consistent with under-ascertainment in GP records. Taken together, the validation results support a pragmatic use of the registry, with it being well-suited to cohort identification and delivery-focused research, while studies centred on early losses, terminations, or precise gestational measures will benefit from enhanced linkage and outcome refinement.

### Contribution, Limitations, and Future Directions

This work adds to the evidence base by demonstrating the feasibility of generating a regional, reproducible pregnancy registry from routine data using open-source, modular methods adaptable across integrated care systems.[2, 3] This is particularly timely given national moves toward federated analytic infrastructure, notably the NHS Secure Data Environment network through the NHS Data for R&D programme, where consistent, portable cohort-creation algorithms are a prerequisite for valid cross-site inference. It included a sizable longitudinal cohort nearing 400,000 patients over a 10-year period. It also clarifies where routine primary care data performs adequately and where persistent limitations remain, thereby informing appropriate analytic use and prioritising infrastructure improvements. Not all elements of this development were optimal. Reliance on coded primary care data inevitably led to missed or misclassified outcomes, and the absence of maternity-specific fields limited detailed validation. In addition, translation of code lists from Read to SNOMED CT was undertaken pragmatically with limited clinical input during development, which may have resulted in omission of relevant concepts or inclusion of less specific codes. Future research should prioritise updating the SNOMED CT codelists and thorough clinical validation. Improvements in linkage and data availability are also likely to materially enhance completeness and accuracy. In particular, incorporating richer maternity data sources (e.g., the Maternity Services Data Set), alongside improved access to estimated delivery dates, would strengthen pregnancy dating, outcome ascertainment, and the interpretability of gestational measures.

## Conclusion

In summary, the Greater Manchester Pregnancy Registry provides a scalable foundation for pregnancy research using linked routine data in a regional setting. The results align with prior evidence regarding the strengths and limitations of pregnancy identification from routine records. Delivery capture is feasible and moderately consistent across sources, while early outcomes and gestational timing remain challenging. By sharing the methods openly, we aim to support cumulative improvement and collaboration in pregnancy research using real-world data.

## Data Availability

This study was approved via the GMCR's secondary uses and research governance process, which involved review against legal, ethical, and information governance criteria. The authors recognise the Greater Manchester Care Record (a partnership of Greater Manchester Health and Social Care Partnership, Health Innovation Manchester and Graphnet Health, on behalf of Greater Manchester localities) in the provision of data required to undertake this work. However, due to the restrictions surrounding this dataset it cannot be shared. All code will be made available.

https://github.com/ChantelleCornett/GMCRegistry

## Acknowledgements

This study was approved via the GMCR’s secondary uses and research governance process, which involved review against legal, ethical, and information governance criteria. The authors recognise the Greater Manchester Care Record (a partnership of Greater Manchester Health and Social Care Partnership, Health Innovation Manchester and Graphnet Health, on behalf of Greater Manchester localities) in the provision of data required to undertake this work. Chantelle Cornett acknowledges the receipt of studentship awards from the Health Data Research UK-The Alan Turing Institute Wellcome PhD Programme in Health Data Science (Grant Ref: 218529/Z/19/Z).

